# Low Gut Ruminococcaceae Levels are Associated with Occurrence of Antibiotic-associated Diarrhea

**DOI:** 10.1101/2021.09.23.21263551

**Authors:** Xiaoqiong Gu, Jean XY Sim, Wei Lin Lee, Liang Cui, Yvonne FZ Chan, Ega Danu Chang, Yii Ean Teh, An-Ni Zhang, Federica Armas, Franciscus Chandra, Hongjie Chen, Shijie Zhao, Zhanyi Lee, Janelle R. Thompson, Eng Eong Ooi, Jenny G. Low, Eric J. Alm, Shirin Kalimuddin

## Abstract

Patients receiving antibiotics often suffer from antibiotic-associated diarrhea (AAD). AAD is of clinical significance as it can result in premature antibiotic discontinuation and suboptimal treatment of infection. The drivers of AAD however, remain poorly understood. We sought to understand if differences in the gut microbiome, both at baseline and during antibiotic administration, would influence the development of AAD. We administered a 3-day course of oral amoxicillin-clavulanate to 30 healthy adult volunteers, and performed a detailed interrogation of their stool microbiome at baseline and up to 4-weeks post antibiotic administration, using 16S rRNA gene sequencing. Lower levels of Ruminococcaceae were significantly and consistently observed from baseline till Day 7 in participants who developed AAD. The probability of AAD could be predicted based on qPCR-derived levels of *Faecalibacterium prausnitzii*, the most dominant species within the Ruminococcaceae family. Overall, participants who developed AAD experienced a greater decrease in microbial diversity during antibiotic dosing. Our findings suggest that a lack of gut Ruminococcaceae at baseline influences development of AAD. In addition, quantification of *F. prausnitzii* in stool prior to antibiotic administration may help identify patients at risk of AAD, and aid clinicians in devising individualised treatment regimens to minimise such adverse effects.

## Introduction

Antibiotic-associated diarrhea (AAD) occurs in a significant proportion of patients, and is particularly associated with use of broad-spectrum antibiotics (1). AAD may sometimes be severe enough to result in premature discontinuation of antibiotics, and can in turn result in suboptimal treatment of infection. AAD has also been shown to prolong hospital stay, increase risk of other infections and lead to higher overall healthcare costs (2). Therefore, AAD is of significant clinical importance, and a better understanding of its underlying mechanisms and drivers is needed in order to devise therapeutic strategies to minimise its occurrence.

While it is well known that antibiotics disrupt and alter the diversity of microorganisms within the gut (gut microbiome), it is less clear how AAD develops (3). One hypothesis is that AAD results from the overgrowth of toxigenic bacteria, such as *Clostridium difficile*, which are resistant to the administered antibiotic (4). However, *C. difficile* diarrhea only accounts for 15-25% of all cases of AAD (5). Other possible pathogens which have been implicated include *Clostridium perfringens, Staphylococcus aureus, Escherichia coli, Pseudomonas aeruginosa*, and *Klebsiella pneumoniae* (3, 6), supported by studies in murine models demonstrating overgrowth of these pathogens in mice with AAD (6, 7). Another proposed mechanism of AAD is the loss of functional and beneficial gut microbes with critical metabolic activities, resulting in reduced carbohydrate fermentation and short-chain fatty acids (SCFAs) that are important for colonic health (8, 9). Despite these mechanistic explanations for AAD, many gaps in knowledge still remain. Firstly, the etiopathogenesis of non-*C. difficile* AAD is poorly defined. Secondly, although much work has been done in animal models to study the link between gut microbiota alteration and AAD (6, 7), studies in humans are lacking. This is of consequence as there are significant differences between the gut microbiome of humans and animals. Thirdly, while post-antibiotic treatment changes could explain AAD through increases of pathogenic bacteria or decreases in beneficial gut microbes, it remains unknown whether the baseline microbiome composition prior to antibiotic administration may confer predisposition to AAD.

We hypothesized that baseline differences in the gut microbiome prior to antibiotic administration could account for why certain patients developed AAD, and that these baseline differences would in turn modulate changes in the gut microbiome composition post antibiotic-administration. To test our hypothesis, we conducted an experimental medicine study in 30 healthy adult volunteers. We administered a 3-day course of oral amoxicillin-clavulanate to study participants. Amoxicillin-clavulanate, a broad-spectrum antibiotic with activity against Gram-positive and Gram-negative organisms, including anaerobes, was chosen as it is one of the most widely-prescribed antibiotics (10), and is associated with a high incidence of AAD (11, 12). We monitored individuals for occurrence of AAD while on amoxicillin-clavulanate. Using 16S rRNA gene sequencing, we tracked dynamic changes in the composition and diversity of the gut microbiome at baseline and up to four-weeks post antibiotic administration, in order to identify differences between individuals who developed AAD (AAD group) and those who did not (non-AAD group).

## results

### Gut Ruminococcaceae levels differentiate AAD and non-AAD groups both at baseline and post-antibiotic treatment

30 healthy adult volunteers were orally administered 1g of amoxicillin-clavulanate twice daily for 3 days, a dose commonly used in clinical practice. Individuals enrolled had a mean age of 30.3 ± 6.2 years and a mean body mass index of 24.8 ± 3.4 kg m^-2^, with an equal male to female ratio (1:1) (Table S1). Faecal samples were collected prior to (day 0, baseline), during (days 1, 2, 3) and after antibiotic treatment (days 7, 14 and 28) (Figure 1A). Nucleic acids were extracted from faecal samples and subjected to 16S rRNA gene sequencing. Using droplet digital PCR, we confirmed that *C. difficile* toxic TcdA and TcdB genes were not detected in any of the faecal samples at baseline and during-antibiotic treatment (days 1-3), ruling out *C. difficile* colitis as a cause of diarrhea (13). For this study, we *a priori* defined AAD as at least 1 episode of Bristol Stool Scale type 6 or 7 on either days 1, 2 or 3 (Figure 1B). Based on this definition, there were 13 individuals who developed AAD (AAD group) and 17 individuals who did not (non-AAD group) (Table S1-2). There were no differences in demographics between the two groups (Table S1). In order to ensure the safety and well-being of the study volunteers, the study protocol mandated that amoxicillin-clavulanate would be discontinued early if an individual experienced 3 or more episodes of watery stool in a 24-hour period - this occurred in 4 of 13 individuals in the AAD group (Table S1, S3). One individual in the non-AAD group developed severe vomiting after a single dose of amoxicillin-clavulanate, resulting in antibiotic discontinuation on day 1. In all 5 individuals however, faecal samples were collected and sequenced as per protocol (Table S1).

**Figure 1.**
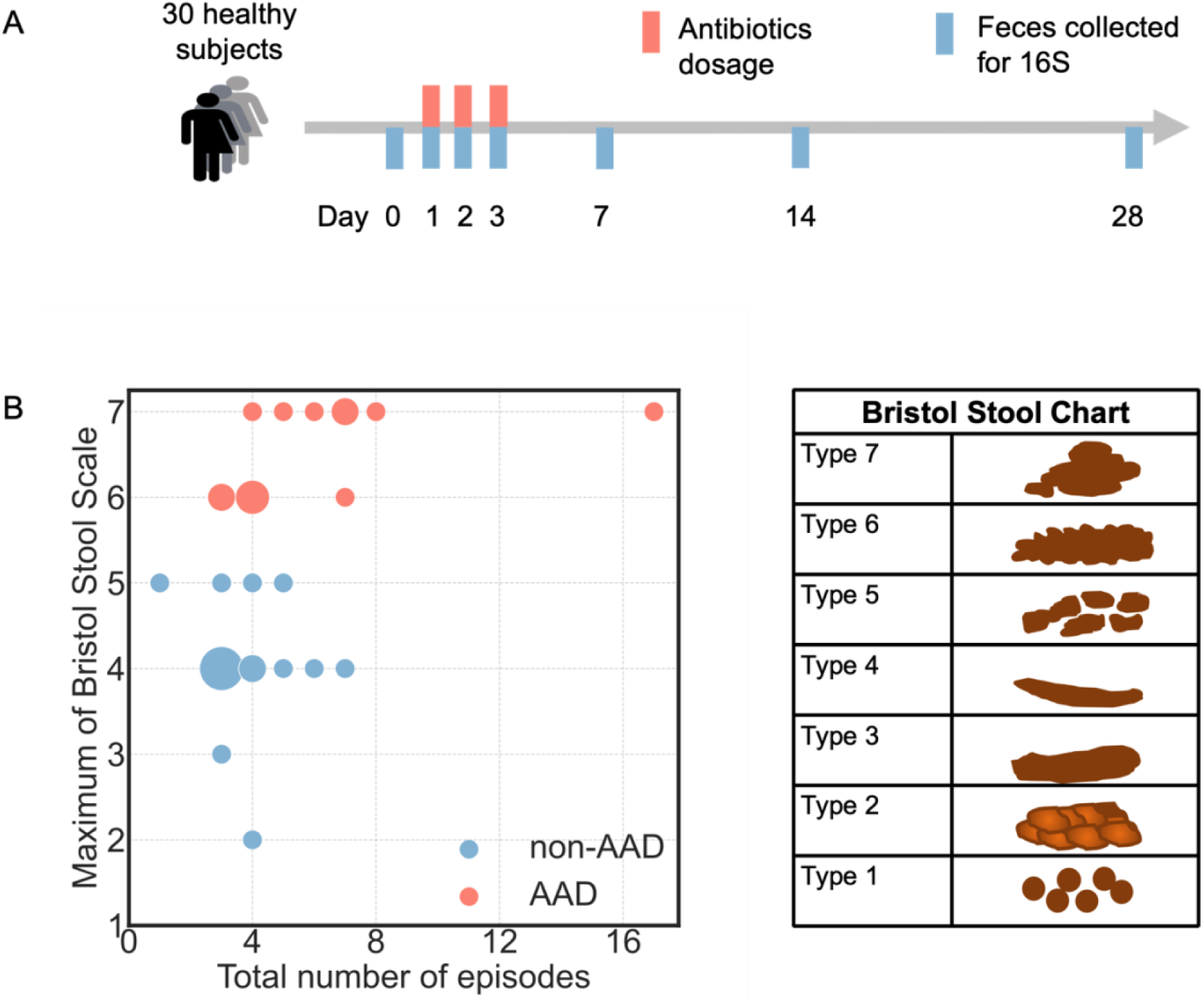
AAD vs non-AAD classification across the duration of the study. (A) Study design of 30 healthy adult volunteers administered amoxicillin-clavulanate with paired faecal sampling. (B) The segregation of AAD and non-AAD groups according to the maximum Bristol Stool Scale on either day 1, 2 or 3 of antibiotic treatment. The total number of episodes indicates the number of episodes across day 1 to 3.

We first examined whether the composition of the gut microbiome at baseline would influence development of AAD. Aggregation of microbial sequence types at the taxonomic levels of “family” and “genus” revealed notable dynamics in the AAD compared to non-AAD groups (Figure 2A, Figure S1A). We observed that Enterobacteriaceae blooms (days 1-3) were more common in the AAD group (76.9% vs 29.4% occurrence frequency) (Figure S1B), with a much higher magnitude (mean 59.1% vs 21.0%, median 67.9% [IQR 33.7-78.7] vs 20.0% [IQR 17.4-25.2], Bonferroni-corrected p < 0.0001) (Figure S1C). Within the Enterobacteriaceae family, these were assigned to the genus Escherichia-Shigella (Figure S1D).

**Figure 2.**
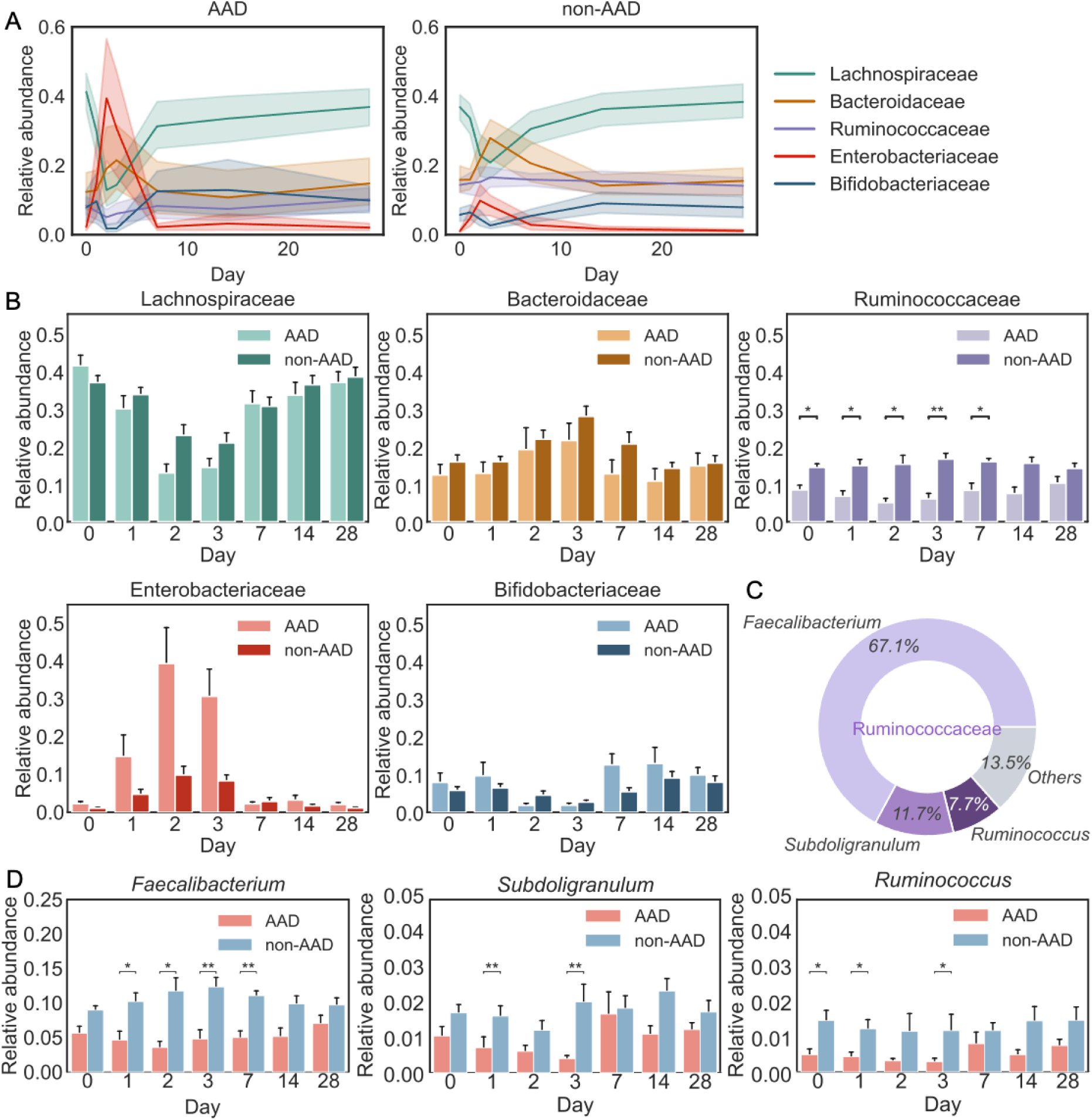
Ruminococcaceae differentiates the AAD and non-AAD groups across the duration of the study. (A) Profound community changes were found in the AAD group compared to the non-AAD group at the family level. Solid lines represent the mean; color shadings represent 95% confidence intervals. (B) Dynamics of 5 most abundant families across the duration of the study between the AAD and non-AAD groups (Bonferroni-corrected, two-sided Mann-Whitney U test, P ≤ 0.05, *; P ≤ 0.01, **). Error bars represent 68% confidence intervals. (C) Pie charts of 3 most abundant genera in Ruminococcaceae across the duration of the study (D) Dynamics of 3 most abundant genera in Ruminococcaceae, *Faecalibacterium, Subdoligranulum* and *Ruminococcus* across the duration of the study between AAD and non-AAD groups (Bonferroni-corrected, two-sided Mann-Whitney U test, P ≤ 0.05, *; P ≤ 0.01, **).

Among the major taxonomies, we looked for features present at baseline that could differentiate between the AAD and non-AAD groups. Significantly, we found that Ruminococcaceae levels were distinctly different between the two groups, both at baseline and post-antibiotic treatment. At baseline, the AAD group had a lower proportion of Ruminococcaceae (mean 8.4% vs 14.4%, median 7.9% [IQR 4.2-11.9] vs 14.2% [IQR 11.6-17.7], Bonferroni-corrected p=0.02, n=30) (Figure 2B). The AAD group also experienced a consistently lower proportion of Ruminococcaceae compared to the non-AAD group till day 7. On average, across the duration of the study, the AAD group had a lower proportion of Ruminococcaceae (mean 7.5% vs 15.3%, median 5.5 [IQR 3.0-10.8] vs 15.6 [IQR 11.8-19.2], Bonferroni-corrected p=2.1e-14, n=197). These differences were not observed with any of the other major taxonomies.

We identified potential drivers of AAD at the genus level within the Ruminococcaceae family across the duration of the study, with *Faecalibacterium* being the most abundant (mean 67.1%, median 66.6 [IQR 56.8-77.7]), followed by *Subdoligranulum* (mean 11.7%, median 10.4 [IQR 6.5-14.5]) and *Ruminococcus* (7.7%, median 6.5 [IQR 3.1-10.1]). We observed that the genera *Faecalibacterium, Subdoligranulum* and *Ruminococcus* were significantly less abundant in the AAD group across most days between day 0 to day 7.

### Amoxicillin-clavulanate causes greater gut microbiome diversity loss and community disturbance in the AAD group compared to the non-AAD group

We next determined the extent to which the gut microbiota was disrupted during antibiotic treatment and the timescale of recovery, in terms of the abundance of microbial amplicon sequence variants (ASVs) and composition. By day 3, the faecal microbiota from the AAD group was distinctly different from microbiomes sampled at the other timepoints and from the non-AAD group, forming a separate cluster along the first and second axes of a principal-coordinate analysis (PCoA) (Figure 3A). We quantified the microbial diversity within each individual at a given time point (α diversity) and the differences between each individual’s baseline and post-treatment gut microbiota (*β* diversity) (Figure 3B). We observed a greater decrease in diversity in the AAD group, compared to the non-AAD group on days 2 and/or 3 (Figure 3B, P < 0.05, Bonferroni-corrected Mann-Whitney U test). Analysis of the relative abundance of bacterial taxonomic groups at the phylum level supported our finding that the AAD group was more severely impacted than the non-AAD group (Figure 3C). This difference in diversity between the two groups was driven by a sharp increase of Proteobacteria, and a decrease of Firmicutes and Actinobacteria in the AAD group, as compared to the non-AAD group on days 2 and 3 (Figure 3C). Individuals in both groups returned to their baseline taxonomy and diversity by day 7, as shown by permutational multivariate analysis of variance (PERMANOVA) (Figure 3A).

**Figure 3.**
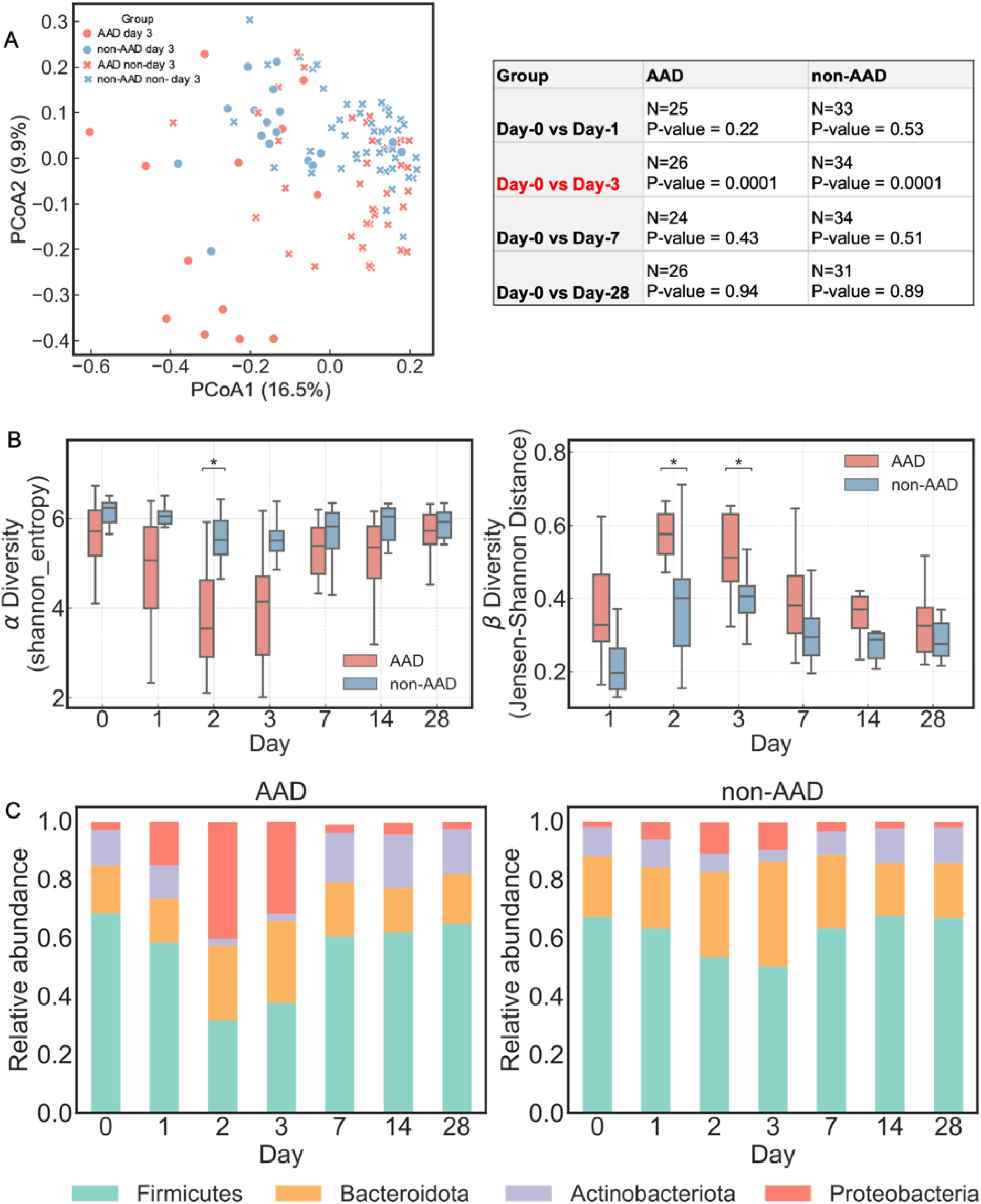
Amoxicillin-clavulanate causes greater gut microbiome diversity loss and community disturbance in the AAD group compared to the non-AAD group. (A) Principal coordinates analysis (PCoA) based on ASV-level Bray-Curtis dissimilarity. Display is based on sample scores on the primary axis (PCoA1, 16.5% variance explained) and secondary axis (PCoA2, 9.9% variance explained). To reduce the redundancy of sample points on the plot, we picked microbiomes on day 3 to represent the post-dosing period (days 1-3). Days 0, 3, 7 and 28 were included as the data points with days 0, 7 and 28 represented simply as ‘non-day 3’. The greatest variation observed in the AAD group occurs on day 3. Individuals return to their baseline microbiomes from day 7. PERMANOVA results show that there were no significant differences between day 0 and 7 in both AAD (P = 0.43, N = 24) and non-AAD groups (P = 0.51, N = 24). (B) Within-sample species diversity (α diversity of ASVs, Shannon entropy index) greatly decreased in the AAD group compared to the non-AAD group on day 2. The similarity of each individual’s gut microbiota to their baseline communities (*β* diversity of ASVs, Jensen–Shannon distance) greatly decreased in the AAD group compared to the non-AAD group cross days 2-3. Significant difference between the AAD and non-AAD groups are labelled with asterisks (Bonferroni-corrected, two-sided Mann–Whitney U test, P ≤ 0.05, *; P ≤ 0.01, **). (C) A sharp increase of Proteobacteria, and a decrease of Firmicutes and Actinobacteria were observed in the AAD group on days 2 and 3.

### Predicting the risk of AAD based on the relative abundance of Ruminococcaceae at baseline

While post-treatment changes in the gut microbiome could explain AAD, predicting which patients are at higher risk of developing AAD would be clinically useful, to enable personalization of antibiotic prescription to minimize the incidence of AAD. Through hierarchical clustering of microbiome composition at baseline, the majority of non-AAD individuals (Figure 4A, blue labels) were grouped into a single cluster (cluster 1), while AAD individuals (Figure 4A, red labels) were separated into various clusters (non-cluster 1). This clustering suggests that some features could be a potential indicator to identify individuals at risk of AAD. Moreover, it also suggests the presence of common characteristics amongst the non-AAD group, but not for the AAD group. Our findings suggest that individuals who went on to develop AAD could be differentiated from those who did not, by the relative abundance of Ruminococcaceae at baseline. This was evident from PCoA based on Bray-Curtis dissimilarities (Figure S2). Hence, we sought to determine if we could predict the risk of AAD based on the relative abundance of Ruminococcaceae at the baseline (D0). Upon ranking the subjects based on their relative-abundance of Ruminococcaceae, we found that there was a clear separation between the AAD and non-AAD groups at the extremes of relative abundance (<0.05 or >0.16) (Figure 4B). Next we quantified absolute values of one species under Ruminococcaceae (*Faecalibacterium prausnitzii* [*F. prausnitzii*], the most abundant Ruminococcaceae species, using a qPCR assay (14). We found that the gene copies of *F. prausnitzii* followed a similar trend with 16S rRNA gene relative abundance of Ruminococcaceae (Spearman’s ρ = 0.85, p = 3.0e-9) (Figure 4C-D). We calculated the risk of developing AAD based on the absolute abundance of *F. prausnitzii* derived using qPCR at baseline. Lower relative abundance of *F. prausnitzii* at baseline were predictive of risk of AAD. The probability of developing AAD was above 0.7 if *F. prausnitzii* levels were less than 2.4×10^7^ GC/μL, and below 0.3 if *F. prausnitzii* levels were above 8.0×10^7^ GC/μL (Figure 4E).

**Figure 4.**
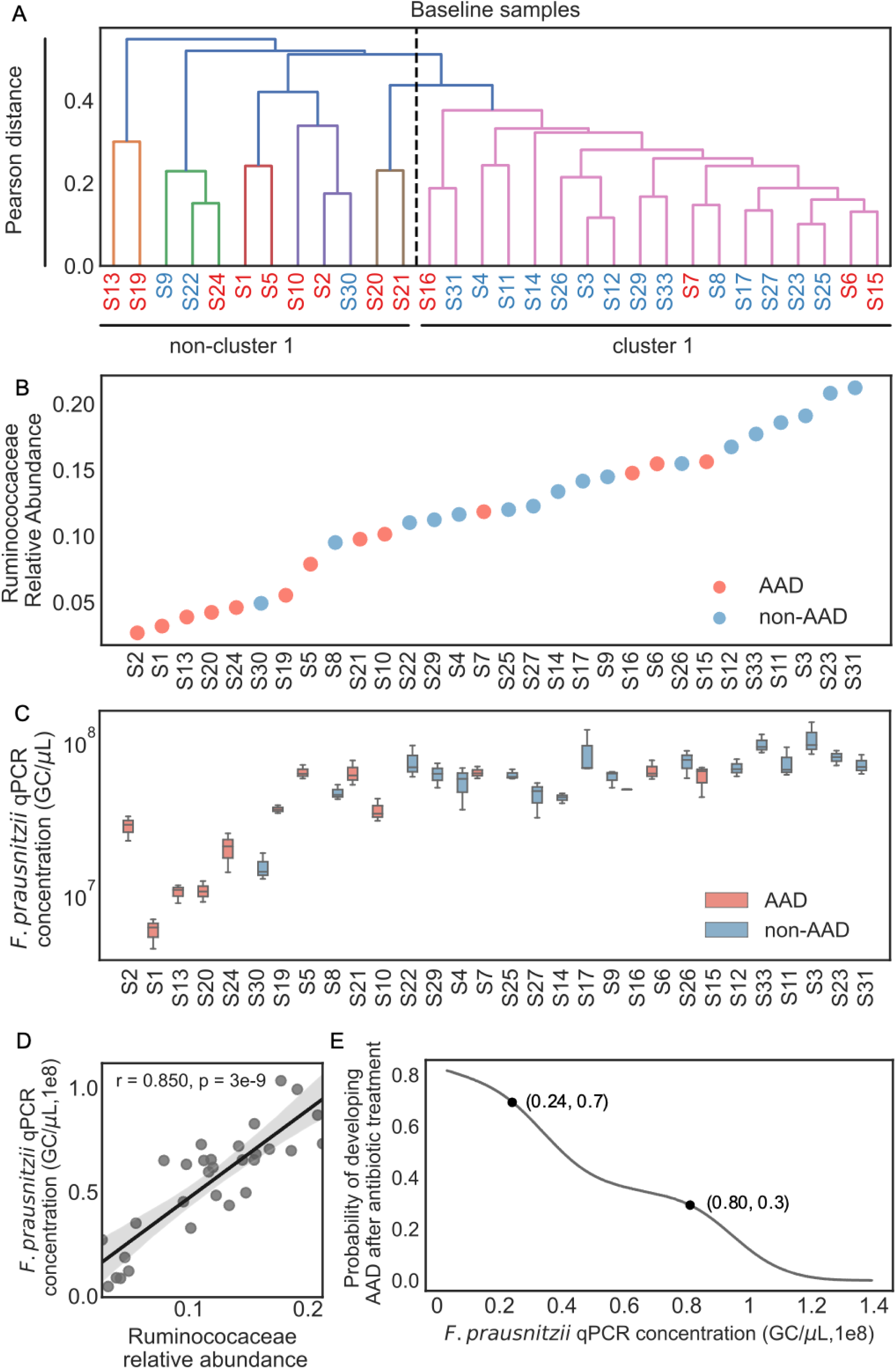
Predicting the risk of AAD based on the relative abundance of Ruminococcaceae at baseline. Red and blue labels denote individuals with AAD and non-AAD, respectively (A) Inter-individual microbial community variation of amplicon sequence variants (ASVs) at baseline. Hierarchical clustering of the baseline faecal bacterial composition of 30 healthy individuals (average linkage, with correlation matrix). The colour threshold for signifying clusters was set to a pearson distance of ‘0.4’. Majority (14/17) of non-AAD individuals were grouped into Cluster 1. Majority of AAD individuals (9/13) were excluded from Cluster 1 to form several individual branches. (B) Distribution of Ruminococcaceae relative abundance at baseline among the AAD and non-AAD groups. Each number refers to the individual at baseline. (C) qPCR concentration for the species *F. prausnitzii* was normalized to 16S rRNA gene copy (Data represent median±IQR range, n=3). Each number refers to the individual at baseline. (D) Correlations between Ruminococcaceae relative abundance and *F. prausnitzii* median absolute quantification (Spearman’s ρ = 0.850, p = 3.0e-9). Line depicts the best linear fit and blue shading the 95% confidence interval of the linear fit. (E) Calculated predictive precision of developing AAD using *F. prausnitzii* absolute abundance quantified by qPCR assay.

## Discussion

Our study has shed important insights on differences in gut microbiome responses between individuals who developed AAD and those who did not. We found that individuals who developed AAD experienced greater gut microbiome community changes, accompanied by lower diversity and a greater disturbance in abundance across taxonomies. Individuals in the AAD group experienced a sharp increase in Proteobacteria, belonging to the genus Escherichia-Shigella. Looking for taxonomic signatures that could differentiate between the two groups, we found that amongst all bacterial families, gut Ruminococcaceae levels were significantly and consistently different between the two groups. Ruminococcaceae levels were lower both at baseline prior to antibiotic treatment, and up till day 7 post-dose. In fact, ranking of gut Ruminococcaceae levels, or simply *F. prausnitzii* (the most dominant species within the Ruminococcaceae family), prior to antibiotic treatment was able to indicate if an individual would develop AAD upon treatment with amoxicillin-clavulanate.

Ruminococcaceae is a group of strictly anaerobic bacteria that is present in the colonic mucosal biofilm of healthy individuals (15). Decreased abundance of Ruminococcaceae has been implicated in a number of inflammatory bowel diseases, including ulcerative colitis and Crohn’s disease (16–18), inflammatory diseases such as hepatic encephalopathy (19) and has also been associated with *C. difficile* infection and *C. difficile*-negative nosocomial diarrhea (20). Ruminococcaceae plays an important role in the maintenance of gut health through its ability to produce butyrate and other SCFAs. These SCFAs are essential carbon and energy sources to colonic enterocytes (21), in the absence of which, functional disorders of the colonic mucosa may occur – which may manifest in the form of osmotic diarrhea (9). Indeed, supplementation of butyrate and other SCFAs has been shown to reduce colonic inflammation and improve diarrhea in conditions such as inflammatory bowel diseases, irritable bowel syndrome, and diverticulitis (22–24). We thus posit that a lack of Ruminococcaceae resulting in decreased SCFA production may be driving the development of AAD in our cohort of otherwise healthy individuals who received amoxicillin-clavulanate.

To date, many others have studied the role of probiotic bacteria such as *Lactobacillus, Bifidobacterium, Clostridium, Bacillus* and *Lactococcus* in the development and prevention of AAD (25). In a systematic review and a meta-analysis, administration of such probiotics were associated with a reduced incidence of AAD (26, 27). However, a clinical trial performed on a cohort of 3000 older patients concluded that *Lactobacillus* and *Bifidobacterium* probiotic administration was not found to be effective in preventing *C. difficile -* associated diarrhea (1). Likewise, *Lactobacillus reuteri* was not effective in preventing AAD in a cohort of 250 children (28, 29). There remains a need to search for probiotic candidates that will be effective against AAD. Our findings suggest that certain species within the Ruminococcaceae family may be useful as probiotics to prevent AAD, but this will need to be further evaluated in randomised-controlled clinical trials.

We have identified clear differences in baseline gut microbial composition that may enable pre-identification of individuals at higher risk of developing AAD. Although our findings at present are only applicable in the context of AAD caused by amoxicillin-clavulanate, our study provides a framework to identify potential drivers of AAD caused by other classes of antibiotics. We acknowledge that a limitation of our study is the relatively small sample size, which may have reduced the statistical power given the variability of inter-individual differences in the gut microbiome. Despite this, we were still able to observe clear and significant differences between the AAD and non-AAD groups.

Our findings provide evidence for the first time that baseline differences in the individual’s gut microbial composition can influence the risk of developing AAD with certain antibiotics, and would guide the development of point-of-care diagnostics. Being able to pre-identify individuals at increased risk of AAD would aid clinicians in devising an individualised antibiotic regime best suited to the patient that is least likely to result in premature antibiotic discontinuation and suboptimal treatment of infection - An example of such a clinical workflow is presented in Figure 5. In addition, the use of Ruminococcaceae as a prebiotic to prevent AAD in patients who receive amoxicillin-clavulanate also warrants further exploration. Overall, our study provides insights into how the gut microbiome influences development of AAD, and opens a window of opportunity for further research in this area.

**Figure 5.**
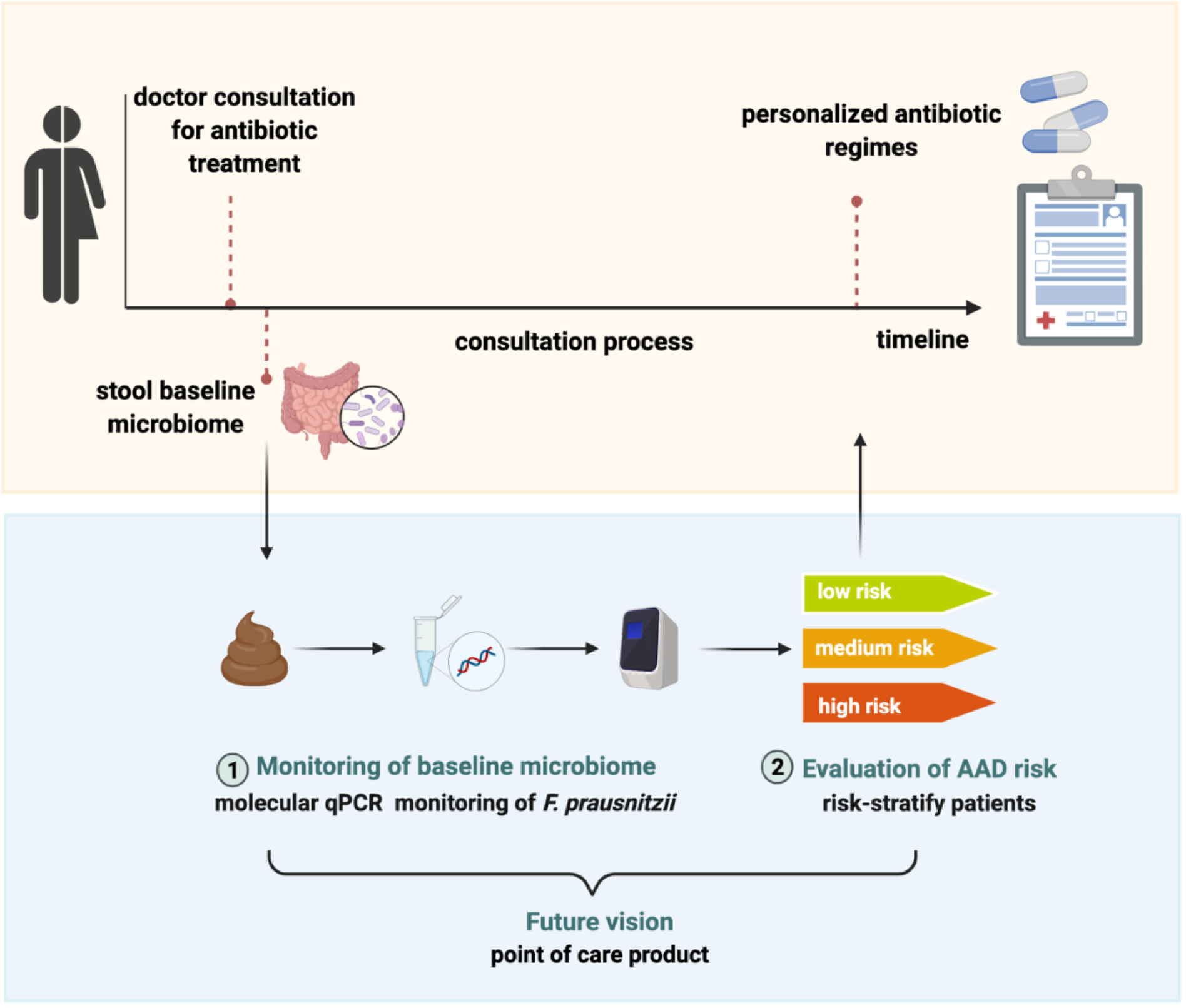
Risk-stratifying patients at risk of AAD using stool F. prausnitzii qPCR. We propose a workflow to evaluate AAD risk at baseline prior to antibiotic administration. This would enable clinicians to risk-stratify patients, and identify those in whom use of amoxicillin-clavulanate should be avoided due to higher risk of AAD. Although quantification of *F. prausnitzii* is currently via qPCR, this could potentially be translated into rapid point-of-care diagnostics in furture. This framework could also serve as basis for identifying potential drivers in AAD caused by other classes of antibiotics. (This figure was created using BioRender.com.)

## Methods

### Study design and participants

From Aug 2019 to Jan 2020, 30 healthy adult volunteers who fulfilled the pre-determined inclusion and exclusion criteria were enrolled into the study. Individuals were eligible for the study if they were: 1) Aged between 21-40 years, 2) Willing and able to provide written informed consent, and 3) Agreeable to abstain from probiotics and/or prebiotics during the study period. Individuals were excluded if they met any of the following criteria: 1) Presence of underlying chronic medical illness, 2) History of *C. difficile* diarrhea, 3) Inflammatory bowel disease or any other chronic gastrointestinal tract illness, 4) Allergy to beta-lactam antibiotics, 5) Acute infection in the preceding 7 days, 6) Were pregnant or breastfeeding, and/or 7) Receipt of antibiotics in the past 3 months.

Upon enrolment, individuals received oral amoxicillin-clavulanate at a dose of 1g (875 mg of amoxicillin trihydrate and 125 mg of potassium clavulanate) twice a day for 3 days, i.e. a total of 6 doses. Study drug compliance was assessed via daily phone calls and pill count at each study visit. Individuals were instructed to maintain consistent dietary habits and abstain from pre- or probiotics throughout the duration of the study.

### Metadata and sample collection

Baseline demographics were recorded. Individuals were provided with a standardised diary to record frequency of bowel opening and any adverse events. Individuals were followed-up for 28 days (screening, baseline [prior to antibiotic administration], and days 1, 2, 3, 7, 14 and 28). At each study visit, frequency of bowel opening in the past 24 hours was recorded and a faecal sample passed on the day of the study visit was collected. Faecal samples were collected prior to (day 0, baseline), during (days 1, 2, 3) and after antibiotic treatment (days 7, 14 and 28), over a time span of 4 weeks. Faecal samples were collected using a disposable commode 2-piece specimen collector (MEDLINE, USA) and stored immediately in −20°C freezers prior to being transported to the laboratory which was off-site. The Bristol Stool Scale of each faecal sample was assessed by the same study team member each time to ensure consistency.

### DNA extraction, library construction and Illumina 16S rRNA Sequencing

DNA was extracted from approximately 300 - 600 mg of faecal samples using DNeasy® PowerSoil® Pro Kit (Qiagen, Germany, Cat# 47016) following the manufacturer’s instructions. DNA purity and quantity of extracted DNA was determined by Nanodrop® Spectrophotometer ND-1000 (Thermo Fisher Scientific, USA) before sending to the NovogeneAIT Genomics Singapore for sequencing. Bacterial 16S V4 region was amplified with the Earth Microbiome Project recommended primer pairs 515F (GTGCCAGCMGCCGCGGTAA) and 806R (GGACTACHVGGGTWTCTAAT) (30) using Phusion® High-Fidelity PCR Master Mix (New England Biolabs, Cat# M0531L). The size of the amplicon was checked using 1 % agarose gel electrophoresis. The amplicon with correct size was purified from agarose gel using Qiagen Gel Extraction Kit (Qiagen, Germany) and proceeded to library preparation using NEBNext Ultra DNA Library Prep Kit for Illumina® (New England Biolabs, Cat# E7370L) following manufacturer’s instructions. Unique indexes were added to each sample. The library was quantified using qPCR. The sequencing libraries were normalized and pooled at equimolar concentration before performed on NovaSeq-6000 (Illumina, USA) to generate 250 bp paired-end raw reads.

### 16S rRNA gene sequencing datasets pre-processing analysis

Paired-end raw reads were assigned to a sample by their unique barcode, and the barcode and primer sequence were then truncated. Paired-end reads were merged using FLASH (V1.2.7) (31) to merge pairs of reads when the original DNA fragments are shorter than twice of the reads length. The obtained splicing sequences were called raw tags. Quality filtering were then performed on the raw tags under specific filtering conditions of QIIME (V1.7.0) (32) quality control process. After filtering, high-quality clean tags were obtained. The tags were compared with the reference database (Gold database, http://drive5.com/uchime/uchime_download.html) using UCHIME algorithm (33) to detect chimeric sequences, and then the chimeric sequences were removed to obtain the Effective Tags finally. Sequence analysis and processing of paired-end demultiplexed sequences were performed in Quantitative Insights into Microbial Ecology pipeline (QIIME 2, v 2020.6) (34). Demultiplexed sequences were imported using QIIME 2 “Fastq manifest” format by mapping the sample identifiers to absolute file paths containing sequence information for each sample. PairedEndFastqManifestPhred33V2 format was used. Interactive quality plot and a summary distribution of sequence qualities at each base pair position in the sequence data was visualized to determine input parameters for denoising. DADA2 was used to denoise, dereplicate and filter chimaeras in paired-end sequences to identify all amplicon sequence variants (ASVs), equivalent to 100% Operational Taxonomy Unit (OTUs) (35, 36). Forward and reverse reads were truncated at 200 bases to retain high quality bases respectively. In total, we characterized an average of 150,087 ± 14,526 (mean ± SD) 16S rRNA sequences for 197 samples.

### Microbiome composition and diversity

The alpha diversity metrics (Shannon entropy) and beta diversity metrics (Jensen-Shannon Distance) from ASVs were generated via q2-diversity plugin. A sampling depth of 96,935 sequences per sample was used and optimal alpha-rarefaction curves were achieved. Taxonomy assignment to ASVs was performed using q2-feature-classifier plugin using a pre-trained Naive Bayes classifier against the reference 515F/806R region of sequences in Silva 138 at 99% OTUs (37). We computed ASV pairwise distances using the Pearson correlation (ASV abundances across 30 subjects at baseline). The resulting distance matrix was subsequently inputted into a hierarchical clustering function (‘fcluster’). The linkage approach was set as ‘average’. The colour threshold was set to ‘0.4’. Principal coordinate analysis (PCoA) was performed using the ‘scipy’ package in python based on the ASV-level Bray-Curtis dissimilarities between the composition of baseline samples. PERMANOVA analysis was calculated using the ‘skbio’ package in python based on the ASV-level Bray-Curtis dissimilarities and 9999 permutations.

### Calculating predictive probability of developing AAD from baseline abundance

To calculate the predictive probability of developing AAD from *F. prausnitzii* baseline abundance, the absolute abundance of *F. prausnitzii* was min-max normalized to their transformed value between 0 and 1. The predictive probability of developing AAD was calculated using kernel density estimation with a Gaussian distribution kernel. We assumed that each sample had an independent and identical distribution with a mean at its concentration and a standard deviation, which is the hyperparameter of this model. We aggregated the distribution of every sample from the AAD group and obtained the AAD probability density function (PDF). The same approach was applied to calculating the non-AAD PDF. The probability of developing AAD was calculated as 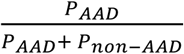.

### Molecular ddPCR assay for C. difficile toxin genes TcdA and TcdB quantification

We excluded *C. difficile* infection in all individuals via PCR of toxin genes TcdA and TcdB in stool at baseline (day 0) and during-antibiotic treatment (days 1-3). Briefly, droplet digital PCR was performed on both 10x and 100x diluted DNA extracted from all baseline and during-antibiotic treatment (days 1-3) samples on Bio-Rad droplet digital PCR system (Bio-Rad, California). Reaction mixtures of 22 μL were prepared with 11 μL of ddPCR supermix for probes (no dUTP) (Bio-Rad, Cat# 1863024), 0.2 μM of forward, reverse primers each, 0.2 μM of fluorescent probes and 2.2 μL of template DNA (13). After that, 20 μL of the reaction mixtures were transferred into the cassette with 70 μL of droplet generator oil for probes (Bio-Rad, Cat# 1863005) for droplet generation. 10,000 - 20,000 droplets were generated from 20 μL of each reaction mix with the QX200 droplet generator (Bio-Rad). Droplet-partitioned samples were transferred to a Twin-tec PCR 96-well plate (Eppendorf), sealed and amplified in the thermal cycler under the Bio-Rad recommended thermal cycling protocol (95°C for 10 min, followed by 40 cycles of 94°C for 30s and 60°C for 1 min, ending with 98°C for 1 min with a ramp rate of 2°C/s). The amplified samples were immediately transferred to the QX200 reader (Bio-Rad) and read in the FAM channel. Analysis of the ddPCR data was performed using QuantaSoft software (Bio-Rad). gBlocks Gene Fragments (Integrated DNA Technologies, Iowa) were designed with DNA sequences corresponding to the amplification regions of the primer-probe sets as positive control (Table S5).

### Molecular qPCR assay for *Faecalibacterium prausnitzii* and 16S rRNA gene characterization

To determine the absolute concentration of *F. prausnitzii* and 16S rRNA gene, qPCR was performed on 100x diluted DNA extracted from all baseline samples (14, 38). Reaction mixtures of 10 μL of extracted stool DNA were prepared in triplicates with 5 μL of qPCR SsoAdvanced Universal SYBR Green Supermix (Biorad, Cat# 1725271), 0.25 μM of forward, reverse primers each and 1 μL of template DNA. The reactions are set up using electronic pipettes (Epperdorf), sealed and amplified in the Bio-Rad CFX384 real-time PCR thermal cycler under the Bio-Rad recommended thermal cycling protocol (98°C for 3 min for polymerase activation and DNA denaturation, followed by 40 cycles of 95°C for 15 s and 60°C for 30 s). All baseline samples for both assays are set up in one 384 plate to avoid inter-plate variation. No template controls were included for both assays. Melt-curve analysis was performed from 65 °C - 95 °C in 0.5 °C increments at 5 sec/step. gBlocks Gene Fragments (Integrated DNA Technologies, Iowa) were designed with DNA sequences corresponding to the amplification regions of the primer-probe sets (Table S5). 10-fold serial dilutions were performed on the gBlocks for a range of 1 to 1 × 10^7^ copies/μL, representing the range of the standard curve for quantification. gBlocks Gene Fragments containing *F. prausnitzii* and 16S rRNA gene were quantified and calibrated using ddPCR to determine the absolute gene copies.

*F. prausnitzii* gene copies per PCR reaction were normalized to the 16S rRNA gene. The standard curve for *F. prausnitzii* was y=37.342-3.674x (efficiency = 85.1%) and for 16S rRNA gene was y=38.230-3.693x (efficiency = 85.4%). To adjust the *F. prausnitzii* for each sample due to nucleic acids extracted from uneven biomass of stool samples, we first calculated the deviation of the 16S rRNA gene from the median of the 16S rRNA gene in all baseline samples, i.e., 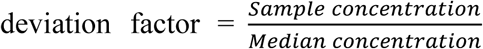. We then divided the *F. prausnitzii* concentrations by this deviation. The final absolute concentration of *F. prausnitzii* (GC/μL per PCR reaction) = The adjusted absolute concentration of *F. prausnitzii* (GC/μL per PCR reaction) * 100 (the dilution factor).

### Quantification and Calibration of gBlocks Gene Fragments

gBlocks Gene Fragments containing F. prausnitzii and 16S rRNA gene were quantified and calibrated using ddPCR to determine the absolute gene copies. Reaction mixture of 22 μL of gBlock Gene Fragments were prepared in duplicates with 11 μL of ddPCR EvaGreen Supermix (Biorad, Cat# 1864034), 0.2 μM of forward, reverse primers each and 2.2 μL of template standard DNA. The reactions are set up in the Bio-Rad CFX96 real-time PCR thermal cycler under the Bio-Rad recommended thermal cycling protocol (95°C for 5 min for polymerase activation, followed by 40 cycles of 95°C for 30 s for DNA denaturation and 60°C for 1 min for annealing/extension, 4°C for 5 min and 90 °C for 5 min for signal stabilization). During the process, the ramp rate was set as 2°C/s.

### Data Availability

The sequencing datasets generated during this study are available at European Nucleotide Archive (ENA): PRJEB46061. Deposited data includes FASTQ files for the 197 16S rRNA amplicon sequences, with adaptors removed and filtered for good quality. The code to reproduce all of the analysis and figures in this paper is available at https://github.com/XiaoqiongGu/augmentin_16S/Final_Figure.

### Statistical analysis

Statistical significance of bacterial diversity and bacterial composition aggregated at ‘family’ and ‘genus’ level between the AAD and non-AAD groups was calculated using two-sided Mann-Whitney U-test. Multiple hypothesis testing in each dataset with Bonferroni correction was used with Mann-whiteney test and ‘two-sided’ method in ‘statannot’ package. All statistical analysis was performed using Python v3.6. The statistical tests used, the definition of statistical significance and multiple tests corrections were indicated in the figure legends.

### Study Approval

This study was approved by the SingHealth Centralised Institutional Review Board (Ref: 2019/2377). Written informed consent was obtained from all participants prior to inclusion in the study.

## Author contributions

Clinical portion of the study and subject enrolment, S.K., J.X.Y.S., Y.E.T. and Y.F.Z.C.; Conceptualization and study design, S.K., E.J.A., E.E.O. and J.G.L.; Stool sample pre-processing, X.G., E.D.C. and L.C.; Molecular qPCR/ddPCR analysis, X.G. and Z.L.; Sequencing data analysis, X.G., E.J.A., W.L.L., F.A., F.C., A.N.Z., H.C. and E.D.C.; Writing-Original Draft, X.G., S.K., W.L.L, E.J.A., F.A. and F.C.; Writing - Review & Editing, X.G., S.K., W.L.L, E.J.A., J.S.X.Y., L.C., F.A., F.C., Y.F.Z.C., A.N.Z., Y.E.T., H.C., S.Z., J.R.T., E.E.O., and J.G.L. All authors agreed to submit the manuscript, read and approved the final draft and take full responsibility of its content, including the accuracy of the data.

## Acknowledgments

We thank the Singapore General Hospital Clinical Trials Research Centre and all study volunteers. We thank the SMART AMR core team (Tse Mien Tan, Peiying Ho, Megan Mcbee, and Farzad Olfat) for their support with logistics and lab supplies; Hafiz Ismail (SMART) for his assistance in quantifying *C. difficile* in the baseline faecal samples; Mathilde Poyet (MIT) and Charmaine Ng (NUS) for their guidance in the stool samples collection and nucleic acids extraction. This study was funded by a SingHealth Academic Medicine Research Grant (AM-CT003-2018) and the National Research Foundation, Singapore, under its Campus for Research Excellence and Technological Enterprise (CREATE) program funding to the Singapore-MIT Alliance for Research and Technology (SMART) Antimicrobial Resistance Interdisciplinary Research Group (AMR IRG).

