## Supporting information for "Low Gut Ruminococcaceae Levels are Associated with Occurrence of Antibiotic-associated Diarrhea"

**Table S1. Demographic Characteristics of Healthy Volunteers.** Subject 18, 28, and 32 did not meet the requirements to participate in the study during screening and were not dosed with antibiotics. Individuals with AAD are highlighted in yellow.

| ID | Age range | Sex | Ethnicity | Height (m) | Weight (kg) | BMI[a] | Total doses of Amoxicillin-Clavulanate |
|---|---|---|---|---|---|---|---|
| 1 | 36-40 | Female | Chinese | 1·61 | 68 | 26·4 | 6 |
| 2 | 26-30 | Male | Malay | 1·68 | 71 | 25·0 | 4[c] |
| 3 | 31-35 | Female | Chinese | 1·61 | 58 | 22·2 | 6 |
| 4 | 36-40 | Male | Chinese | 1·79 | 68 | 21·1 | 6 |
| 5 | 26-30 | Male | Chinese | 1·77 | 96 | 30·5 | 6 |
| 6 | 36-40 | Male | Chinese | 1·77 | 82 | 26·1 | 6 |
| 7 | 36-40 | Female | Chinese | 1·71 | 73 | 24·9 | 6 |
| 8 | 26-30 | Female | Chinese | 1·63 | 69 | 25·8 | 6 |
| 9 | 21-25 | Female | Chinese | 1·62 | 52 | 20·0 | 6 |
| 10 | 21-25 | Male | Chinese | 1·67 | 67 | 24·1 | 6 |
| 11 | 21-25 | Female | Chinese | 1·65 | 52 | 19·1 | 6 |
| 12 | 26-30 | Female | Chinese | 1·56 | 58 | 23·8 | 6 |
| 13 | 31-35 | Female | Chinese | 1·65 | 64 | 23·4 | 6 |
| 14 | 36-40 | Female | Chinese | 1·58 | 61 | 24·4 | 6 |
| 15 | 21-25 | Female | Chinese | 1·63 | 48 | 18·1 | 1[c] |
| 16 | 26-30 | Male | Chinese | 1·6 | 74 | 28·9 | 5[c] |
| 17 | 36-40 | Female | Chinese | 1·59 | 74 | 29·4 | 6 |
| 19 | 21-25 | Male | Chinese | 1·8 | 88 | 27·2 | 6 |
| 20 | 36-40 | Female | Chinese | 1·58 | 57 | 22·7 | 6 |
| 21 | 31-35 | Male | Malay | 1·69 | 68 | 23·9 | 6 |
| 22 | 21-25 | Female | Chinese | 1·60 | 74 | 28·9 | 6 |
| 23 | 31-35 | Male | Chinese | 1·78 | 108 | 34·1 | 6 |
| 24 | 26-30 | Male | Chinese | 1·71 | 73 | 24·8 | 3[c] |
| 25 | 21-25 | Male | Chinese | 1·53 | 50 | 21·7 | 1[b] |
| 26 | 21-25 | Male | Chinese | 1·75 | 77 | 25·0 | 6 |
| 27 | 21-25 | Male | Chinese | 1·72 | 69 | 23·3 | 6 |
| 29 | 21-25 | Male | Chinese | 1·77 | 70 | 22·4 | 6 |
| 30 | 36-40 | Male | Chinese | 1·76 | 78 | 25·3 | 6 |
| 31 | 26-30 | Female | Indian | 1·64 | 73 | 27·1 | 6 |
| 33 | 26-30 | Female | Vietnamese | 1·51 | 52 | 22·9 | 6 |

[b]Subject 25 developed severe vomiting after a single dose of amoxicillin-clavulanate, resulting in discontinuation of antibiotics on day 1.
[c]Subject 2, 15, 16 and 24 experienced 3 or more episodes of watery stool in a 24-hour period (Table S2), resulting in discontinuation of antibiotics.

**Table S2. Demographic Characteristics of Individuals Between the AAD And Non-AAD Groups.** Two-tailed Mann-Whitney tests were performed to test for significant differences between the AAD and non-AAD groups.

| | AAD individuals (n=13) | non-AAD individuals (n=17) | p value |
|---|---|---|---|
| Sex | -- | -- | 0·46 |
| Female | 5(38%) | 10(59%) | |
| Male | 8(62%) | 7(41%) | |
| Age | 30(26-37) | 28(24-34) | 0·34 |
| BMI | 24.86(23.91-26.40) | 23.75(22.19-25.84) | 0·39 |
| Height | 1.68(1.63-1.71) | 1.63(1.59-1.75) | 0·31 |
| Weight | 70.65(67.10-74.00) | 68.65(57.8-73.95) | 0·46 |

Data are n/N (%) for proportional data and median (IQR) for continuous data, unless specified otherwise.

**Table S3. Frequency and Bristol Stool Scale type.** Faeces samples collection log book from 20 Aug 2019 to 20 Jan 2020. Subject 18, 28 and 32 failed during screening and were not dosed with antibiotics. Individuals with AAD are highlighted in yellow. Gray shaded boxes indicate that donors could not produce the stool samples or the stool samples were not collected. A total of 197 faeces samples were collected**.**

| | Screening | | Day 0 | | Day 1 | | Day 2 | | Day 3 | | Day 7 | | Day 14 +/- 2 | | Day 28 +/- 2 | |
|---|---|---|---|---|---|---|---|---|---|---|---|---|---|---|---|---|
| ID | BS | Frequency | BS | Frequency | BS | Frequency | BS | Frequency | BS | Frequency | BS | Frequency | BS | Frequency | BS | Frequency |
| 1 | 4 | 2 | 4 | 2 | 5 | 3 | 6 | 2 | 6 | 2 | 4 | 2 | 4 | 1 | 4 | 2 |
| 2 | 3 | 3 | 3 | 3 | 3 | 4 | 5 | 5[a] | 7 | 8 | NA | NA | 3 | 4 | 4 | 3 |
| 3 | 4 | 1 | 4 | 1 | 4 | 1 | 4 | 1 | 4 | 2 | 4 | 2 | 4 | 1 | 4 | 2 |
| 4 | 3 | 1 | 3 | 1 | 3 | 1 | 3 | 1 | 4 | 1 | 3 | 1 | 4 | 1 | NA | 0 |
| 5 | 5 | 1 | 5 | 3 | 7 | 2 | 5 | 1 | 6 | 2 | 4 | 1 | 4 | 1 | 4 | 3 |
| 6 | 4 | 2 | 5 | 2 | 6 | 2 | 6 | 1 | 6 | 1 | 4 | 1 | 4 | 1 | 1 | 1 |
| 7 | 4 | 1 | 4 | 1 | 6 | 2 | 6 | 2 | 5 | 1 | 4 | 1 | 4 | 2 | 2 | 1 |
| 8 | 4 | 2 | 4 | 1 | 5 | 1 | 5 | 1 | 5 | 1 | 4 | 1 | 3 | 2 | 4 | 2 |
| 9 | 4 | 1 | 4 | 1 | 4 | 1 | 5 | 1 | 5 | 2 | 4 | 1 | 3 | 1 | 4 | 2 |
| 10 | 4 | 2 | 4 | 1 | 4 | 1 | 4 | 1 | 7 | 4 | NA | NA | NA | NA | 4 | 4 |
| 11 | 3 | 1 | 4 | 1 | 4 | 1 | 4 | 2 | 4 | 1 | 2 | 1 | 3 | 1 | 4 | 1 |
| 12 | 4 | 1 | 4 | 1 | NA | NA | NA | NA | 4 | 1 | 4 | 1 | 4 | 1 | 4 | 1 |
| 13 | 5 | 1 | 6 | 1 | 6 | 1 | 5 | 1 | 4 | 2 | 6 | 2 | 4 | 1 | 6 | 2 |
| 14 | 3 | 1 | 4 | 1 | 4 | 1 | 4 | 2 | 4 | 1 | 4 | 2 | 4 | 2 | 3 | 1 |
| 15 | 3 | 1 | 3 | 1 | 7 | 3[b] | 6 | 3 | 4 | 1 | 5 | 1 | 4 | 2 | 3 | 1 |
| 16 | 4 | 1 | 6 | 1 | 6 | 1 | 6 | 2 | 7 | 5[c] | 6 | 1 | 6 | 2 | 6 | 1 |
| 17 | 4 | 1 | 4 | 2 | 4 | 1 | 4 | 1 | 4 | 1 | 4 | 1 | 4 | 1 | 4 | 1 |
| 19 | 6 | 1 | 3 | 2 | 7 | 2 | 2 | 1 | 3 | 1 | 2 | 1 | 4 | 1 | 3 | 1 |
| 20 | 3 | 1 | 4 | 1 | 6 | 1 | 6 | 1 | 3 | 1 | 3 | 1 | 3 | 1 | 4 | 2 |
| 21 | 2 | 1 | 4 | 1 | 4 | 2 | 6 | 1 | 3 | 1 | 4 | 1 | 3 | 2 | 6 | 1 |
| 22 | 5 | 1 | 6 | 1 | 4 | 1 | 4 | 1 | 4 | 1 | 4 | 1 | 4 | 1 | 4 | 2 |
| 23 | 4 | 1 | 3 | 2 | 4 | 1 | 4 | 1 | 4 | 1 | 4 | 2 | 4 | 1 | 4 | 2 |
| 24 | 5 | 2 | 5 | 1 | 7 | 3[d] | 4 | 2 | 1 | 2 | 4 | 2 | 4 | 1 | 4 | 2 |

| | | | | | | | | | | | | | | | | |
|---|---|---|---|---|---|---|---|---|---|---|---|---|---|---|---|---|
| 25 | 3 | 1 | 3 | 1 | 4 | 1 | 5 | 1 | 4 | 1 | 5 | 1 | 4 | 1 | 4 | 1 |
| 26 | 4 | 1 | 3 | 2 | 3 | 1 | 4 | 2 | 4 | 2 | 4 | 2 | 4 | 2 | 4 | 2 |
| 27 | 4 | 1 | 4 | 2 | 5 | 1 | 5 | 3 | 5 | 1 | 4 | 1 | 4 | 2 | 4 | 1 |
| 29 | 4 | 1 | 4 | 2 | 4 | 2 | 4 | 3 | 4 | 2 | 5 | 2 | 4 | 2 | 4 | 3 |
| 30 | 4 | 1 | 4 | 2 | 2 | 1 | 2 | 2 | 2 | 1 | 5 | 4 | 4 | 1 | 4 | 1 |
| 31 | 4 | 3 | 4 | 4 | 4 | 2 | 4 | 2 | 4 | 2 | 4 | 2 | 5 | 1 | 4 | 2 |
| 33 | 2 | 1 | 1 | 2 | 2 | 1 | 3 | 1 | 2 | 1 | 1 | 1 | NA | NA | NA | NA |

[a]Subject 2 experienced 5 episodes of watery stool in a 24-hour period, resulting in discontinuation of antibiotics on day 2, with the total doses of amoxicillin-clavulanate as 4.
[b]Subject 15 experienced 3 episodes of watery stool in a 24-hour period, resulting in discontinuation of antibiotics on day 1, with the total doses of amoxicillin-clavulanate as 1.
[c]Subject 16 experienced 5 episodes of watery stool in a 24-hour period, resulting in discontinuation of antibiotics on day 3, with the total doses of amoxicillin-clavulanate as 5.
[d]Subject 24 experienced 3 episodes of watery stool in a 24-hour period, resulting in discontinuation of antibiotics on day 1, with the total doses of amoxicillin-clavulanate as 3.

**Table S4. Primer-probe sets used in this study**

| Name | Sequences | References |
|---|---|---|
| 16S_V4_515F primer | GTGCCAGCMGCCGCGGTAA | (1) |
| 16S_V4_806R primer | GGACTACHVGGGTWTCTAAT | |
| F. prau_FP | GATGGCCTCGCGTCCGATTAG | (2) |
| F. prau_RP | CCGAAGACCTTCTTCCTCC | |
| 16S_FP | ACTCCTACGGGAGGCAGCAG | (3) |
| 16S_RP | ATTACCGCGGCTGCTGG | |
| TcdA_FP | CAGTCGGATTGCAAGTAATTGACAAT | (4) |
| TcdA_RP | AGTAGTATCTACTACCATTAACAGTCTGC | |
| TcdA_Probe | /56-FAM/TTGAGATGATAGCAGTGTCAGGATTG/36-TAMSp/ | |
| TcdB_FP | TACAAACAGGTGTATTTAGTACAGAAGATGGA | |
| TcdB_RP | CACCTATTTGATTTAGMCCTTTAAAAGC | |
| TcdB_Probe | /56-FAM/TTTKCCAGTAAAATCAATTGCTTC/36-TAMSp/ | |

**Table S5. gBlocks Gene Fragments sequences for *Faecalibacterium prausnitzii*,16S rRNA, *C. difficile* TcdA and TcdB gene.**

| gBlocks Gene Fragments | Sequences |
|---|---|
| gBlocks Gene Fragments for *Faecalibacterium prausnitzii* and 16S rRNA positive control | GGTAGAGGGAAAAGGAGCAATCCGCTTTGAGATGGCCTCGCGTCCGATTAGCTAGTTGGTGA GGTAATGGCCCACCAAGGCGACGATCGGTAGCCGGACTGAGAGGTTGAACGGCCACATTGG GACTGAGACACGGCCCAGACTCCTACGGGAGGCAGCAGTGGGGAATATTGCACAATGGGGG AAACCCTGATGCAGCGACGCCGCGTGGAGGAAGAAGGTCTTCGGATTGTAAACTCCTGTTGT TGAGGAAGATAATGACGGTACTCAACAAGGAAGTGACGGCTAACTACGTGCCAGCAGCCGC GGTAATACGTAGGTCACAAGCGTTGTCCGGAATTAC |
| gBlocks Gene Fragments for *C. difficile* TcdA positive control | GGTAGAGGGAAAAGGAGCAATCCGCTTTGAGATGGCCTCGCGTCCGATTAGCTAGTTGGTGA GGTAATGGCCCACCAAGGCGACGATCGGTAGCCGGACTGAGAGGTTGAACGGCCACATTGG GACTGAGACACGGCCCAGACTCCTACGGGAGGCAGCAGTGGGGAATATTGCACAATGGGGG AAACCCTGATGCAGCGACGCCGCGTGGAGGAAGAAGGTCTTCGGATTGTAAACTCCTGTTGT TGAGGAAGATAATGACGGTACTCAACAAGGAAGTGACGGCTAACTACGTGCCAGCAGCCGC GGTAATACGTAGGTCACAAGCGTTGTCCGGAATTAC |
| gBlocks Gene Fragments for *C. difficile* TcdB positive control | TACACCCCTGCGGCAACGTTGAAGCTCCTGGATTACACTGGCTGGATCTAAGCCGTGACACC CGTCATACTCCATAACCGTCTGTAACTCACGGCTTGTTCTGGACTGGATTGCCATTCTCTCAG AGTATTATGCAGGCCGGCGTACGGGTCCCATATAAACCTGTCATAGCTTACCTGACTCTACTT GGAAATGTGGCTTACAAACAGGTGTATTTAGTACAGAAGATGGATTTAAATATTTTGCCCCA GCTAATACACTTGATGAAAACCTAGAAGGAGAAGCAATTGATTTTACTGGAAAATTAATTAT TGACGAAAATATTTATTATTTTGATGATAATTATAGAGGAGCTGTAGAATGGAAAGAATTAG ATGGTGAAATGCACTATTTTAGCCCAGAAACAGGTAAAGCTTTTAAAGGTCTAAATCAAATA GGTG |

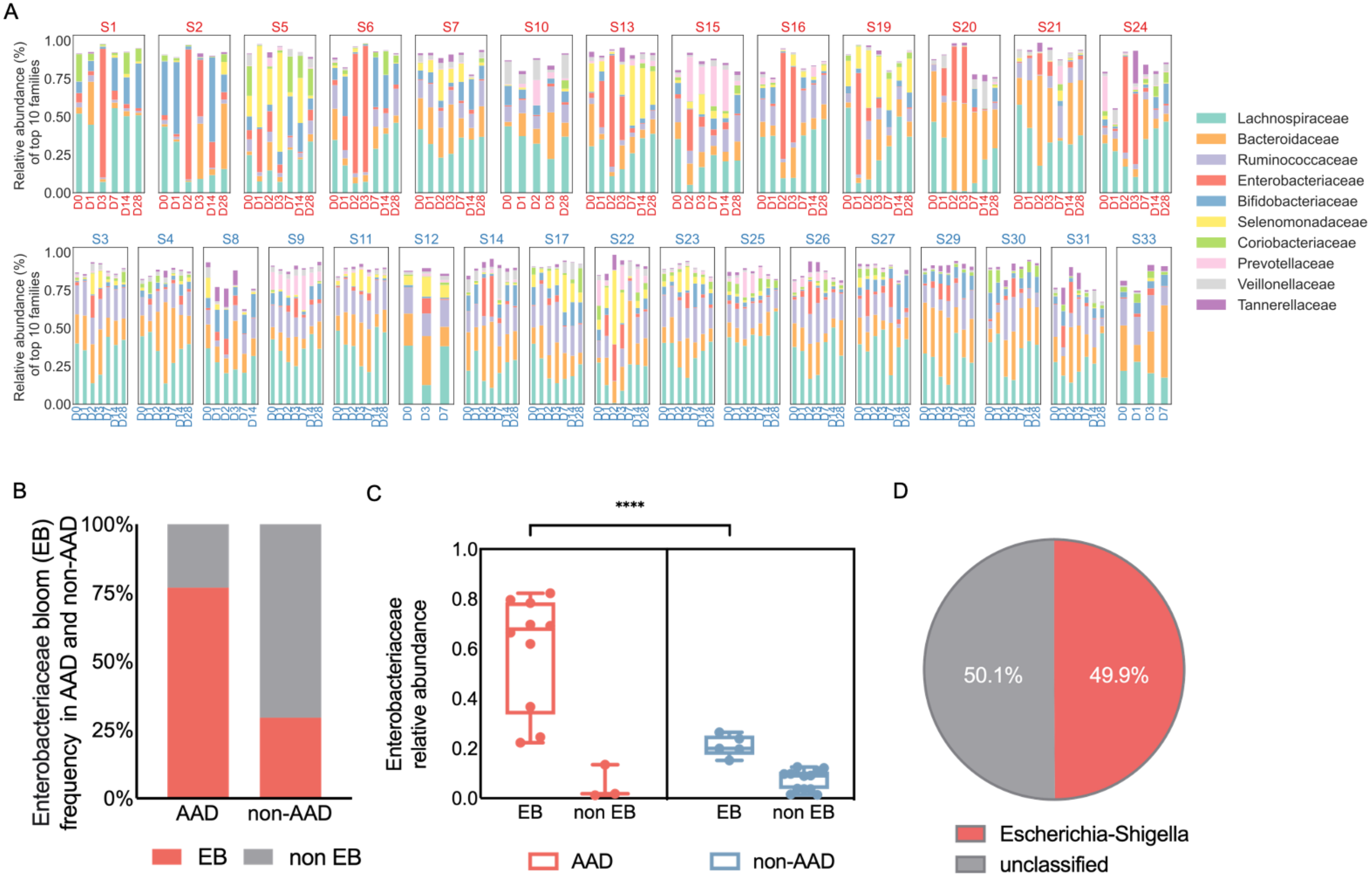

A
S1 S2 S5 S6 S7 S10 S13 S15 S16 S19 S20 S21 S24
S3 S4 S8 S9 S11 S12 S14 S17 S22 S23 S25 S26 S27 S29 S30 S31 S33
Relative abundance (%) of top 10 families
1.00 0.75 0.50 0.25 0.00
Lachnospiraceae
Bacteroidaceae
Ruminococcaceae
Enterobacteriaceae
Bifidobacteriaceae
Selenomonadaceae
Coriobacteriaceae
Prevotellaceae
Veillonellaceae
Tannerellaceae
B
Enterobacteriaceae bloom (EB) frequency in AAD and non-AAD
100% 75% 50% 25% 0%
AAD
non-AAD
EB
non EB
C
****
Enterobacteriaceae relative abundance
1.0 0.8 0.6 0.4 0.2 0.0
EB
non EB
EB
non EB
AAD
non-AAD
D
50.1%
49.9%
Escherichia-Shigella
unclassified

**Figure S1. Enterobacteriaceae bloom (EB) were more common in the AAD group with a much higher magnitude.**
Related to Figure 2. (A) A panel list of the 10 most abundant families in individuals in the AAD (top panel) and non-AAD groups (bottom panel). (B) Enterobacteriaceae blooms (EB) were observed more frequently in the AAD group compared to the non-AAD group (76·9% vs 29·4%). As the increase in Enterobacteriaceae varies between days 1-3 across different individuals, we defined Enterobacteriaceae blooms as having the maximum of Enterobacteriaceae abundance across days 1-3 in each individual being larger than the median(max(days 1-3)-baseline). (C) For all the individuals who experienced EB, the relative abundance of Enterobacteriaceae is higher in the individuals with AAD compared to the individuals with non-AAD (mean 0·59 vs 0·21, FDR corrected $p < 0{\cdot}0001$). (D) Pie charts of the most abundant genera in Enterobacteriaceae across days 1-3. Within the Enterobacteriaceae family, we observed a higher abundance of Escherichia-Shigella (49·9%) at the genus level.

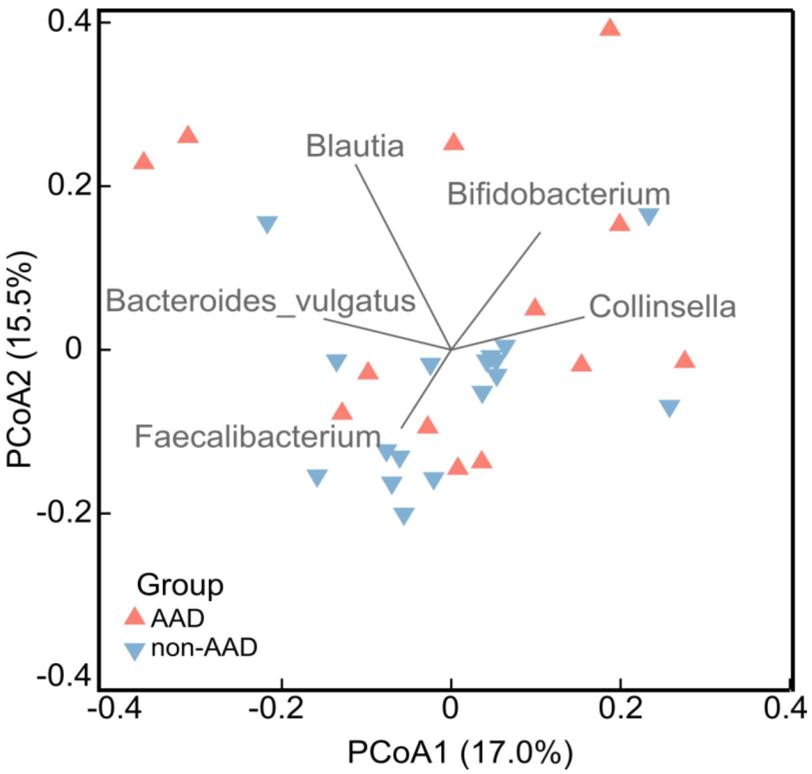

**Figure S2. PCoA of baseline samples based on the ASV level bray-curtis dissimilarity.**
Related to Figure 3. Display is based on sample scores on the primary axis (PCoA1, 17·0% variance explained) and secondary axis (PCoA2, 15·5% variance explained). Vector loading plots at the ASV level indicate that ASV assigned to *Faecalibacterium* points to the non-AAD group while many ununified features (e.g., ASVs assigned to Blautia, Bfidobacterium, Collinsella, Bacteroides vulgatus) point to the AAD groups (multiple linear correlation with the threshold setting as 0·3).
